# Potential causal links between genetic variants in *SAMM50*, *SUGP1*, *MAU2*, and *GATAD2A* and liver fat in individuals with normal weight

**DOI:** 10.1101/2024.11.05.24316758

**Authors:** Ignazio S. Piras, Janith Don, Nicholas J. Schork, Johanna K. DiStefano

## Abstract

**Background & Aims:** Metabolic-associated steatotic liver disease (MASLD) affects individuals across a range of body weights, but little is known about the genetic determinants of this condition in individuals with normal BMI. This study aimed to identify genetic variants contributing to hepatic fat levels in normal-weight individuals.

**Methods:** We conducted a genome-wide association study (GWAS) using magnetic resonance imaging-proton density fat fraction (MRI-PDFF) data from 10,918 normal-weight participants (BMI < 25 kg/m²) of European ancestry from the UK Biobank. We employed a case-control (CC) design to assess hepatic steatosis and a quantitative trait (QT) design for liver fat content. Fine mapping was conducted to identify potential causal variants, while Multi-marker Analysis of GenoMic Annotation (MAGMA) and transcriptome-wide association studies (TWAS) were performed to assess gene-level associations and impute liver-specific gene expression.

**Results:** We identified 241 (CC-GWAS) and 418 (QT-GWAS) significant variants located predominantly in genes on chromosomes 19 and 22, including *PNPLA3*, *TM6SF2*, and *SAMM50*. Fine mapping revealed three potential causal variants in *SUGP1*, *GATAD2A*, and *MAU2*. MAGMA analysis identified eight genes in CC-GWAS and 19 in QT-GWAS, including the novel association with *RFXANK*. TWAS detected *MBOAT7* and *SAMM50*, with *SAMM50* being further implicated as a potential causal gene through TWAS fine mapping.

**Conclusions:** This study, only the second to identify associations at the genome-wide level, represents a significant step forward in understanding the genetic basis of hepatic steatosis in normal-weight individuals, identifying both novel and established genetic variants. These findings provide insights into the pathophysiology of MASLD in this understudied population, emphasizing the importance of genetic predisposition beyond obesity-related mechanisms, and provide potential targets for personalized treatment and prevention strategies.

**Impact and implications:** This study provides relevant insights into the genetic risk factors of MASLD in normal-weight individuals, a population often overlooked in liver disease research. By identifying both novel and established genetic variants, this work highlights the genetic predisposition to hepatic steatosis independent of obesity-related factors. The findings offer potential targets for early detection and therapeutic intervention in normal-weight individuals, which could be applied by physicians, patients, and policymakers to inform precision medicine approaches and preventive strategies. The study provides valuable insights based on a population of European ancestry, and future research can build upon these findings to explore their applicability to patients from diverse ethnic groups.

## Introduction

Obesity is an important risk factor for the development of metabolic dysfunction-associated steatotic liver disease (MASLD) ^1^. MASLD patients with obesity are more likely to develop metabolic dysfunction-associated steatohepatitis (MASH) —often accompanied by hepatic fibrosis— and experience greater liver-related mortality than MASLD patients with overweight ^2,3^. While MASLD prevalence increases in parallel with BMI ^3^, the relationship between obesity and MASLD is complex, as many individuals with obesity maintain normal intrahepatic triglyceride (IHTG) content and metabolic function ^2^, even following moderate weight gain ^4^. Excessive IHTG content in individuals with obesity serves as a strong indicator of metabolic abnormalities, including hepatic, skeletal muscle, and adipose tissue insulin resistance, and dysfunctional free fatty acid metabolism, independent of BMI, percent body fat, and visceral fat mass ^2^. In contrast, normal IHTG content may be protective against the development of obesity-related metabolic complications ^2^. In individuals with normal weight, IHTG content is associated with metabolic dysfunction ^5^. However, in these individuals, hepatic steatosis can be present without insulin resistance, type 2 diabetes, or related metabolic comorbidities ^6^, indicating that excessive liver fat accumulation is not solely dependent on adiposity and can occur in individuals without accompanying metabolic abnormalities.

Numerous studies have compared the characteristics of MASLD among individuals belonging to obese (BMI ≥30 kg/m^2^/ BMI ≥25 kg/m^2^ for Asians), non-obese (BMI <30kg/m^2^/ BMI< 25 kg/m^2^ for Asians), and normal (BMI <25 kg/m^2^/ BMI< 23 kg/m^2^ for Asians) BMI categories ^7-9^. Despite some overlap in clinical presentation among BMI groups, individuals with normal BMI generally exhibit a more favorable metabolic profile, less severe liver disease, and slower disease progression. The characteristics associated with MASLD patients with a normal BMI differ from those observed in individuals with a BMI ≥ 25 kg/m² ^10^, suggesting a complex relationship between adiposity and disease presentation.

Genetic factors likely mediate the development of hepatic steatosis in individuals with normal weight. The *PNPLA3* rs738409 variant has been extensively studied in normal weight MASLD patients ^11–13^. Normal weight carriers of the *PNPLA3* rs738409 GG genotype exhibited the highest MASLD risk compared to individuals with overweight or obesity ^11^, and the impact of rs738409 on MASLD prevalence was more pronounced in individuals with normal weight compared to those with overweight ^14^. Some studies have reported no significant differences in *PNPLA3* genotypes among MASLD patients with normal BMI and those in other BMI classes ^15–17^. Variants in other genes have also been associated with MASLD in individuals within normal BMI ^18–22^.

Despite the evidence linking specific variants to MASLD in individuals with normal weight, the investigation of genetic factors in this population remains limited. Most genetic studies investigating this condition have focused on candidate genes, which are limited by small effect sizes when considered in a genome-wide context and often lack proper adjustment for population structure. To date, only two genome-wide association studies (GWAS) in normal weight individuals have been reported, with only one demonstrating statistically significant results at the genome-wide level ^23,24^. Considering the disparities in MASLD prevalence and risk factors between individuals with normal weight and those with overweight or obesity, we hypothesized that distinct genetic variants might contribute to the susceptibility of MASLD in individuals with a normal BMI. Our objective was to conduct a GWAS to identify genetic variants associated with liver fat specifically in individuals belonging to the normal BMI category. To achieve this, we analyzed magnetic resonance imaging-proton density fat fraction (MRI-PDFF) as both a discrete and quantitative trait, in a cohort of 10,918 individuals with European ancestry in the normal BMI category from the United Kingdom Biobank (UKB). We also performed post-GWAS analyses, including fine mapping to identify potential causal variants, derivation of gene-level statistics (MAGMA), and a transcriptome-wide association study (TWAS) to identify associations between gene expression and hepatic fat levels. Through this multi-analysis approach, we sought to identify and characterize specific genetic variants that contribute to hepatic fat accumulation in individuals with normal weight.

## Methods

A comprehensive description of the methods employed in this study can be found in the supplementary materials.

### Study sample

We used data from the UKB, which contains information from ∼500,000 participants aged between 40 and 69 years recruited from the UK between 2006 and 2010 ^25^. The UKB includes health data, results from physical examinations, and biological samples for genetic analysis.

### Data quality control

An overview of the analytical workflow is shown in **Fig 1**. The UKB imputed genotyped data version 3 was utilized for this study, and preprocessing, quality control, and genetic association analysis were performed using PLINK 2.0. Only variants with high imputation quality (imputed information score > 0.8) and an autosomal location were included. Samples with 1) heterozygosity outside of three standard deviations, 2) kinship value ≥ 0.125, and 3) discrepancies between UKB genetic and self-reported sex were excluded. To establish a homogenous ethnic group, we selected individuals with a self-reported ancestral background of ‘White British’, choosing ‘Caucasians’ from the genetic ethnic grouping. Outliers were removed using principal components and only samples with unambiguous values for the phenotype and covariates used in the association analysis were included. We selected individuals with available MRI-PDFF values up until the date of the analysis, and removed participants with conditions that might impact liver fat levels based on ICD9 and ICD10 codes (**Table S1**). After selecting participants within the normal BMI category (BMI < 25 kg/m^2^), we achieved a sample size of 10,934 individuals.

**Fig. 1.**
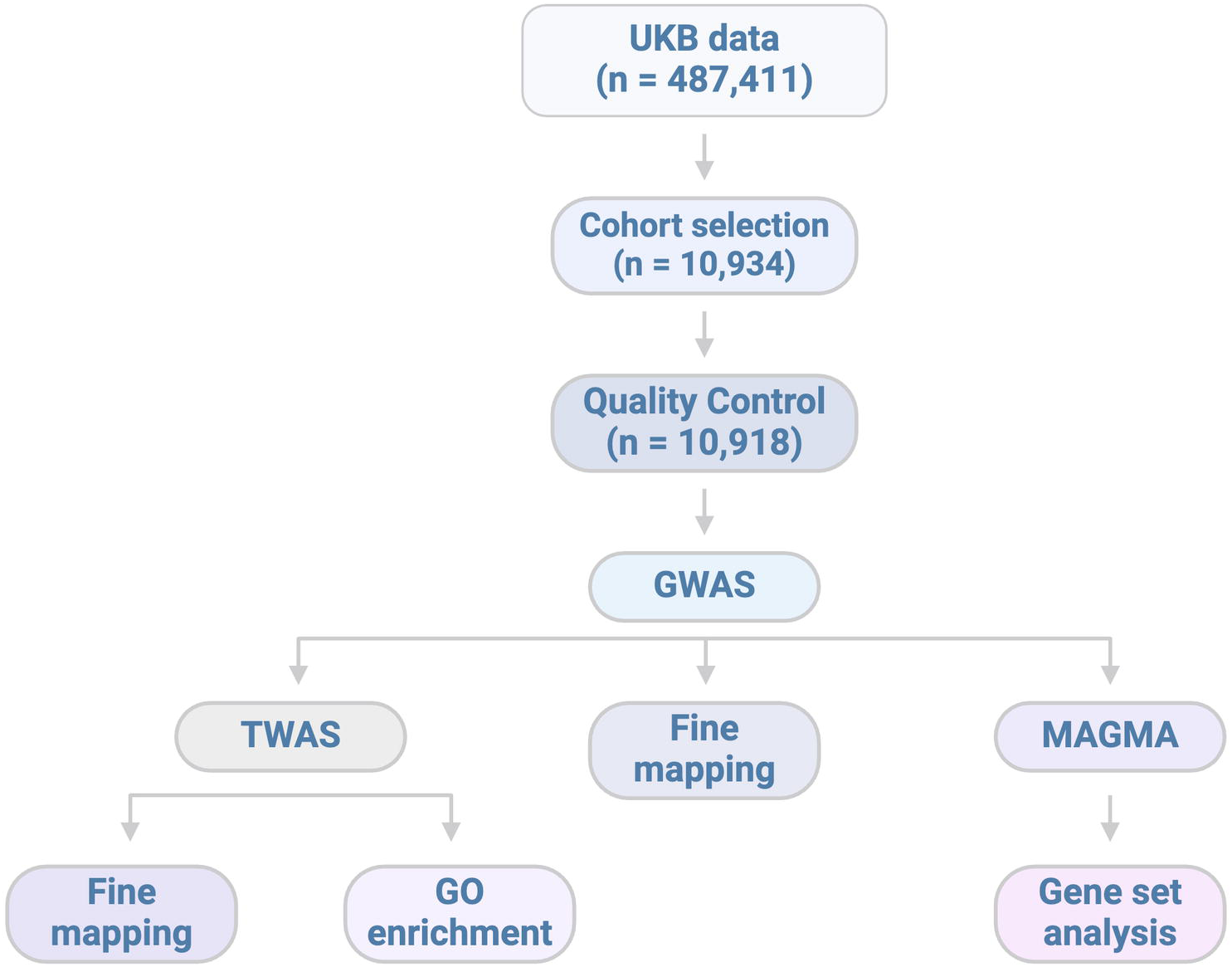
The analytical workflow and study design. The UKB imputed genotyped data version 3, encompassing 487,411 individuals and more than 96 million variants based on the GRCh37 genome build, was utilized for this study. Subsequent filtering and processing steps produced a final cohort comprising 10,918 individuals for the GWAS. The additional analyses following the GWAS were performed as described in the Methods section.

We then excluded variants with a minor allele frequency ≤ 0.05, missingness per individual ≥ 0.05, missingness per marker ≥ 0.05, and Hardy-Weinberg equilibrium p-value ≤ 1.0E-06, obtaining a final sample size of 10,918 individuals and a pool of 6,156,170 variants. The principal component analysis plot for this final dataset is shown in **Fig S1**.

#### GWAS

We conducted two distinct GWAS: a case-control (CC) GWAS, where MRI-PDFF values were used to define individuals with (≥ 5%) or without (< 5%) hepatic steatosis, and a quantitative (QT) GWAS, using the range of MRI-PDFF values. Both GWAS were adjusted for birth year, sex, BMI, alcohol intake frequency (UKB field 1558), and the first 10 genetic principal components (UKB field 22009). To account for multiple testing, we applied the genome-wide cutoff of p < 5.0E-08. Following both analyses, the ‘clump’ function was used to extract variants with a p < 5.0E-08 and variants in linkage disequilibrium (LD) with the most significant variants were removed.

#### GWAS fine-mapping

Fine-mapping analysis was conducted on the significant SNPs using *FINEMAP v.1.4*, with the goal of identifying the causal variants. The algorithm (*stochastic statistic search* method) explores a set of the most probable causal configurations of the region. We considered a posterior inclusion probability (PIP) > 80% as strong evidence of causality, a PIP ≥ 50% as moderate evidence, and a PIP < 50% as weak evidence of causal association between variant and trait.

#### MAGMA

We applied the Multi-marker Analysis of GenoMic Annotation (MAGMA) method, which provides gene-level statistics using a multiple regression approach, to incorporate LD information between markers. The statistic was generated considering 10 kb regions surrounding the gene boundaries. P-values were adjusted for multiple testing using the Bonferroni method, accounting for the number of genes tested. Results were investigated for Gene Ontology enrichment through the MAGMA gene-set analysis.

#### TWAS

Liver expression data models from the Genotype-Tissue Expression Project version 8 (GTEx v8) was used to impute gene expression values, which quantify the relationship between individual genotype and corresponding gene expression levels, thereby capturing *cis*-acting genetic effects. TWAS p-values were adjusted using the Bonferroni method based on the total number of genes included in the GTEx v8 reference (p < 1.34E-05; alpha = 0.05). Joint and conditional tests were conducted for all genes with suggestive adjusted p-value (p < 2.68E-05; alpha = 0.10), to assess whether the signal in genome-wide significant genes was independent of variants located in nearby loci. To identify causal genes associated with the trait, we performed fine mapping using the FOCUS method.

## Results

### Genetic variants on chromosomes 19 and 22 are associated with hepatic steatosis in individuals with normal weight

Out of a total of 10,934 participants, 16 were excluded due to discrepancies between the filtered genetic data and associated covariates. The unfiltered dataset comprised 28,733,793 variants. From this dataset, 580,051 were excluded due to incomplete genotype data, 5,355 for Hardy-Weinberg disequilibrium, and 21,992,217 failed to meet the established MAF cutoff. In total, 6,156,170 common variants were retained for the GWAS analysis.

The CC-GWAS was conducted using 815 cases and 10,103 controls. Characteristics of these study participants are presented in **Table 1**. Participants exhibiting elevated hepatic fat levels were significantly older, had a significantly higher BMI, and were more likely to be male when compared to those with normal liver fat levels. We identified 241 significant SNPs surpassing the genome-wide significance threshold of p < 5.0E-08 with minimal test statistics inflation (**Fig S2A**). These SNPs were clustered within two specific chromosomal regions on 19p12 and 22q13 (**Fig 2A**, **Table 2**, and **Table S2**) and distributed across 16 distinct genes within a range of genomic elements, including exons, introns, and untranslated regions (UTRs). On chromosome 19, the associated region spanned 689,559 base pairs (bp) ranging from 19,103,986 to 19,793,545 and harbored 15 genes. While the rs58542926 variant within the *TM6SF2* gene showed the strongest evidence for association in this locus, the regions exhibiting the highest significance were in closer proximity to the *SUGP1* and *HAPLN4* genes (**Fig S3A**).

**Fig. 2.**
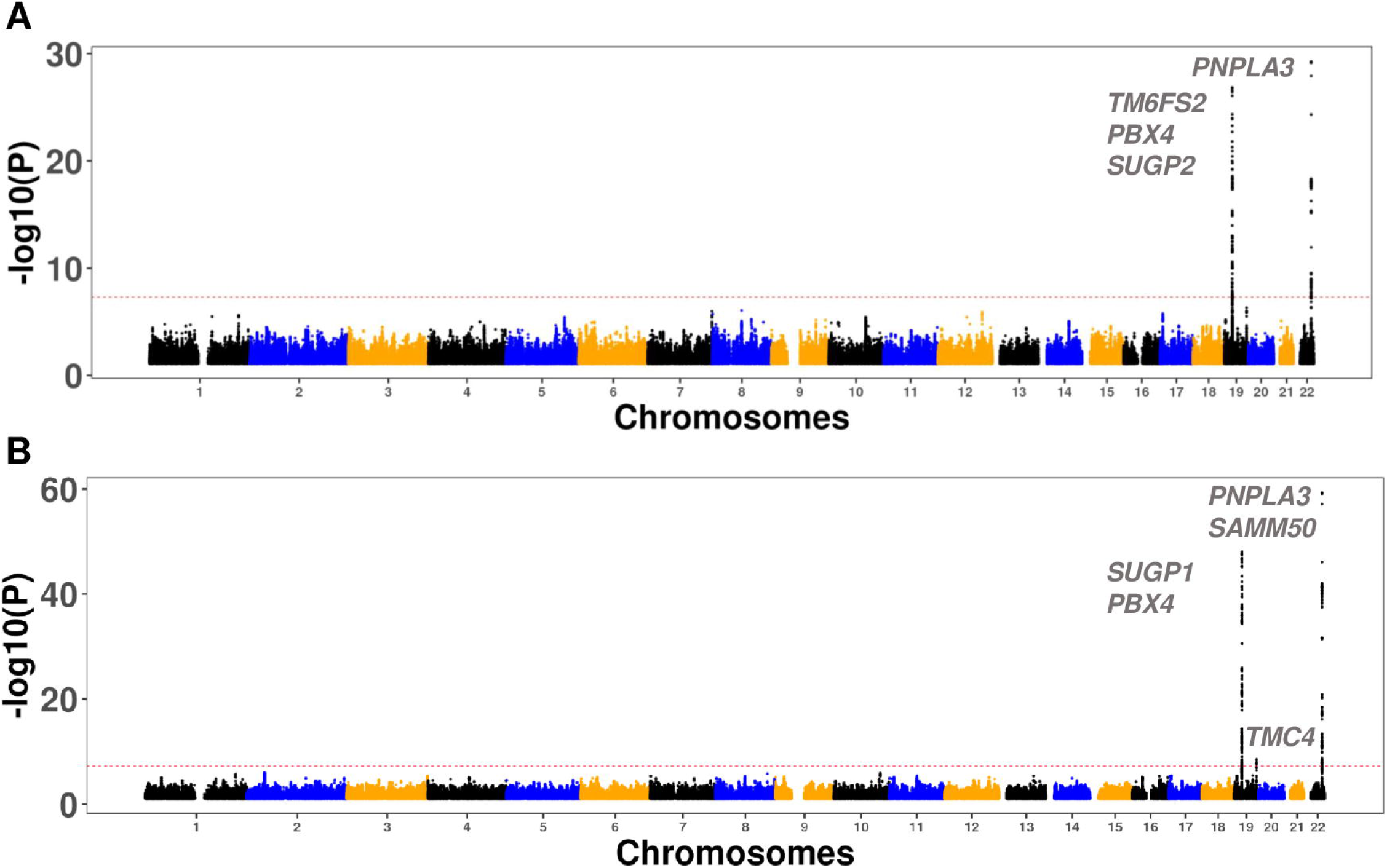
Manhattan plot showing the results for the (A) case-control GWAS (CC-GWAS) and the quantitative GWAS (QT-GWAS). The genes harboring variants associated with liver fat are shown for each analysis. P-values were obtained using a logistic regression (CC-GWAS) and linear regression (QT-GWAS), adjusting for sex, birth year, and the top ten principal components. The red line indicates the genome-wide significance level (p < 5.0E-08).

**Table 1:**
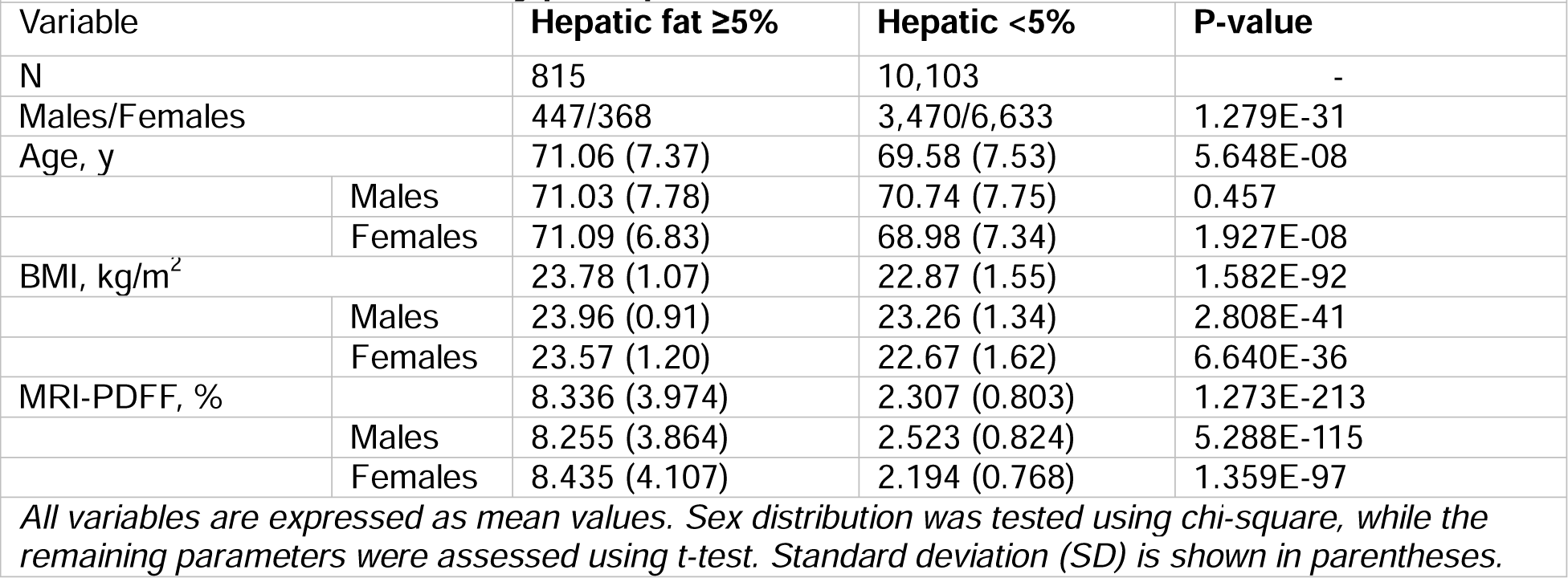
Characteristics of study participants in the CC-GWAS.

| **Table 1: Characteristics of study participants in the CC- GWAS** |  |  |  |  |
| --- | --- | --- | --- | --- |
| Variable | | Hepatic fat $\geq 5\%$ | Hepatic fat $< 5\%$ | P-value |
| N |  | 815 | 10,103 | - |
| Males/Females |  | 447/368 | 3,470/6,633 | 1.279E-31 |
| Age, y |  | 71.06 (7.37) | 69.58 (7.53) | 5.648E-08 |
|  | Males | 71.03 (7.78) | 70.74 (7.75) | 0.457 |
|  | Females | 71.09 (6.83) | 68.98 (7.34) | 1.927E-08 |
| BMI, kg/m<sup>2</sup> |  | 23.78 (1.07) | 22.87 (1.55) | 1.582E-92 |
|  | Males | 23.96 (0.91) | 23.26 (1.34) | 2.808E-41 |
|  | Females | 23.57 (1.20) | 22.67 (1.62) | 6.640E-36 |
| MRI-PDFF, % |  | 8.336 (3.974) | 2.307 (0.803) | 1.273E-213 |
|  | Males | 8.255 (3.864) | 2.523 (0.824) | 5.288E-115 |
|  | Females | 8.435 (4.107) | 2.194 (0.768) | 1.359E-97 |
| *All variables are expressed as mean values. Sex distribution was tested using chi-square, while the remaining parameters were assessed using t-test. Standard deviation (SD) is shown in parentheses.* |  |  |  |  |

**Table 2:** Variants significantly associated with hepatic fat in the CC-GWAS*.

|  |  |  |  |  |  |  |  |  |  |
| --- | --- | --- | --- | --- | --- | --- | --- | --- | --- |
| Variant | Gene | Chrom | Position | Effect allele | Case_Freq | Ctl_Freq | OR | SE | P-value |
| rs738408 | PNPLA3 | 22 | 44324730 | T | 0.335 | 0.212 | 1.92 | 0.057 | 5.35E-30 |
| rs58542926 | TM6SF2 | 19 | 19379549 | T | 0.145 | 0.069 | 2.37 | 0.079 | 1.48E-27 |
| rs17217098 | PBX4 | 19 | 19702384 | A | 0.126 | 0.063 | 2.22 | 0.084 | 1.22E-21 |
| rs73006914 | SUGP2 | 19 | 19110422 | T | 0.081 | 0.049 | 1.80 | 0.101 | 4.97E-09 |
| *\*After LD-clumping* |  |  |  |  |  |  |  |  |  |

On chromosome 22, the region of association extended over 72,586 bp, from 44,324,558 to 44,397,144. This segment included genes such as *PNPLA3*, *SAMM50*, and *PARVB*. Of these, the SNP rs738408 in the *PNPLA3* gene exhibited the most significant association, with a p-value = 5.4E-30 (**Fig S3B**).

Characteristics of the study participants and summary values of the variables used in the QT-GWAS are shown in **Table 3**. We identified 418 significant SNPs predominantly clustered within three regions – two located on chromosome 19 and one on chromosome 22 – and distributed across 24 distinct genes (**Fig 2B**, **Table 4, Table S3**). Like the CC-GWAS, low inflation was observed in this analysis (**Fig S2B**). The first region on chromosome 19 spanned 689,559 bp (positions19,103,986-19,793,545; **Fig 3A**) and was also detected in the CC-GWAS analysis. Within this region, rs200210321, located in the *SUGP1* gene, exhibited the strongest evidence of association. The second region on chromosome 19, which was not identified in the CC-GWAS, spanned 5,768 bp (positions 54,671,421-54,677,189; **Fig 3B**), with the strongest association observed for rs60204587 in the *TMC4* (transmembrane channel like 4) gene.

**Fig. 3.**
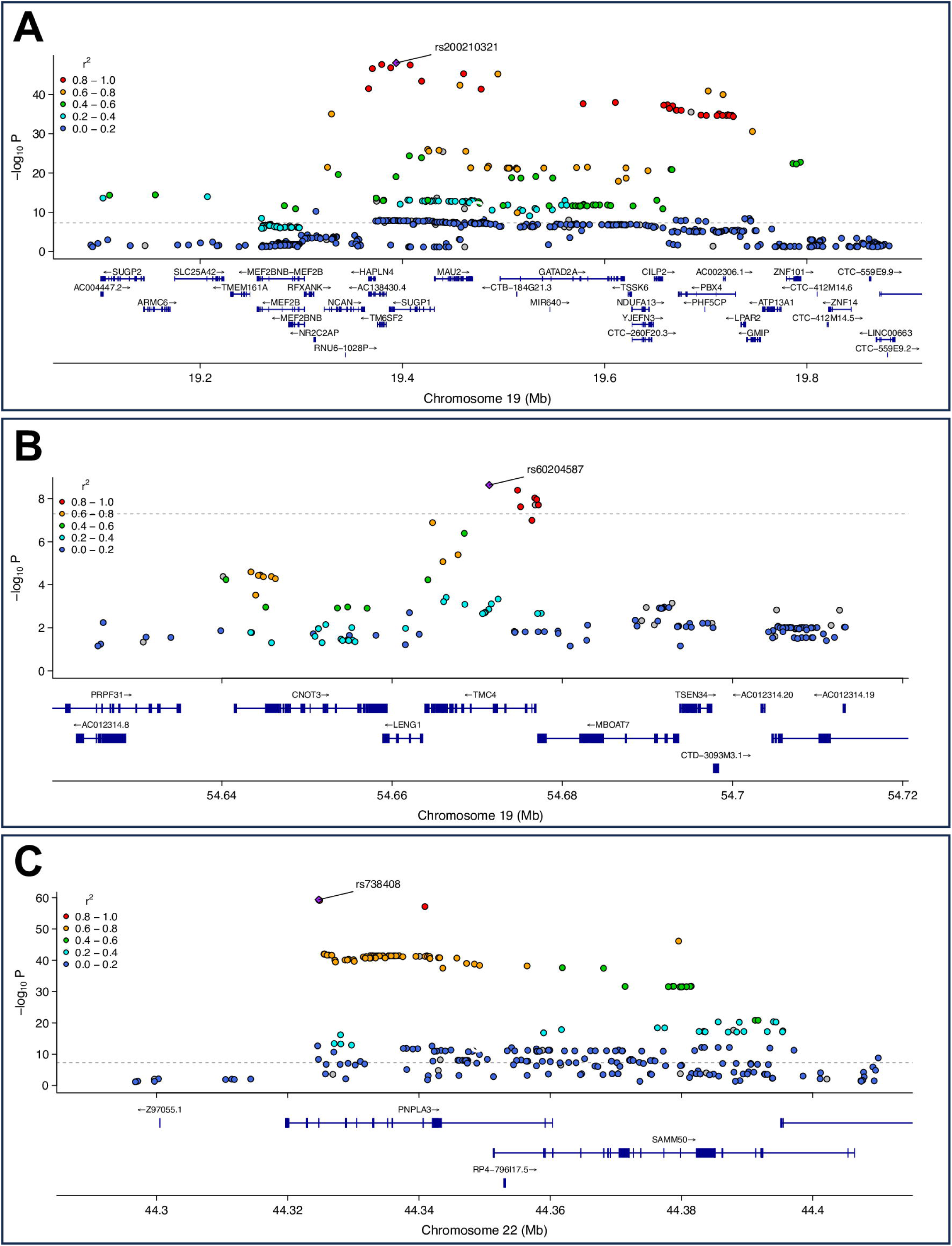
Regional plot showing the three significant loci from the QT-GWAS (p < 5.0E-08). (A) Region 1 on 19p13.11 (positions 19,103,986-19,793,545); (B) Region 2 on 19q13.4 (positions 54,671,421-54,677,189); (C) Region 3 on 22q13.31 (positions 44,324,558-44,409,993). For each region, the gene containing the most significant variant is depicted. P-values were obtained using a linear regression (QT-GWAS), adjusting for sex, birth year, and the top ten principal components. LD was estimated using PLINK2 (*--r* command).

**Table 3:**
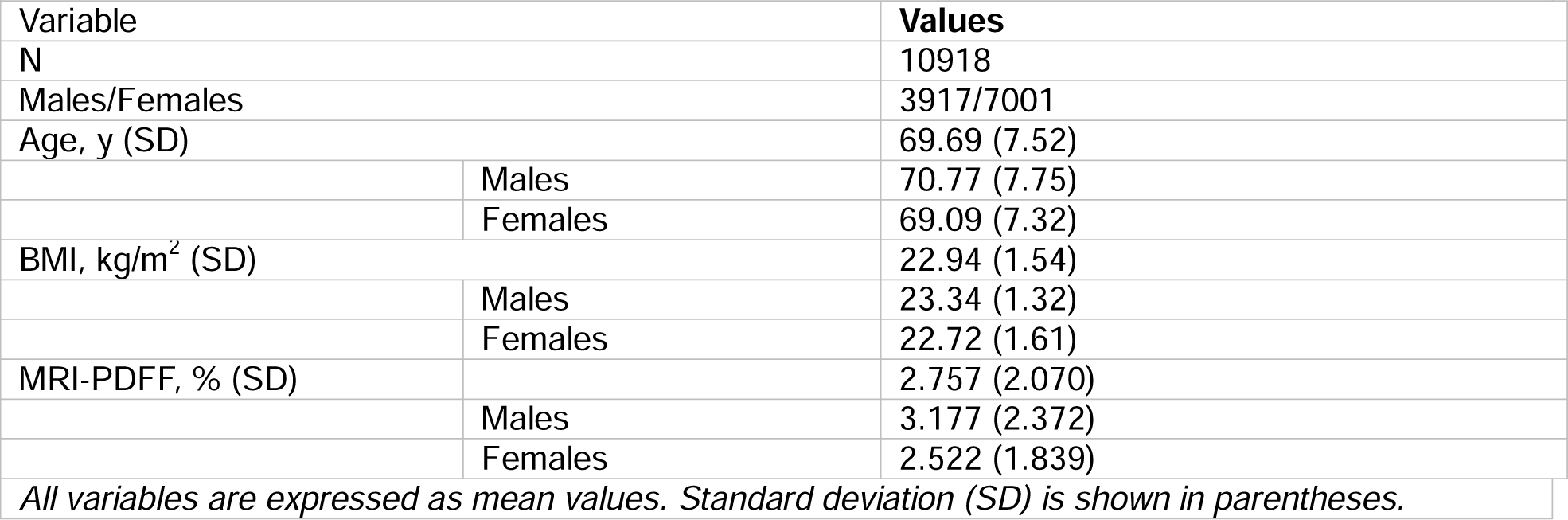
Characteristics of study participants in QT-GWAS.

**Table 4.** Variants significantly associated with hepatic fat in the QT-GWAS*.

|  |  |  |  |  |  |  |  |
| --- | --- | --- | --- | --- | --- | --- | --- |
| **Variant** | **Gene** | **Chrom** | **Position<sup>1</sup>** | **Effect allele** | **BETA** | **SE** | **P-value** |
| rs738408 | *PNPLA3* | 22 | 44324730 | T | 0.532 | 0.032 | 4.31E-60 |
| rs200210321 | *SUGP1* | 19 | 19393890 | AG | 0.766 | 0.052 | 8.92E-49 |
| rs17217098 | *PBX4* | 19 | 19702384 | A | 0.727 | 0.054 | 1.24E-41 |
| rs73006914 | *SUGP2* | 19 | 19110422 | T | 0.487 | 0.062 | 4.64E-15 |
| rs11912828 | *PNPLA3*;<br>*SAMM50* | 22 | 44348116 | A | -0.211 | 0.031 | 1.19E-11 |
| rs12484530 | *PARVB* | 22 | 44409993 | A | 0.292 | 0.048 | 1.61E-09 |
| rs60204587 | *TMC4* | 19 | 54671421 | A | 0.167 | 0.028 | 2.32E-09 |
| rs12609436 | *GMIP* | 19 | 19743098 | T | 0.161 | 0.027 | 3.93E-09 |
| *\*After LD-clumping* |  |  |  |  |  |  |  |

On chromosome 22, the region of association spanned 85,435 bp (positions 44,324,558-44,409,993; **Fig 3C**), and the most significant SNP in this region was rs738408, located in the *PNPLA3* gene. This region was also identified in the CC-GWAS. Overall, 241 SNPs overlapped between the two analyses, with the QT-GWAS capturing all those identified in the CC-GWAS and detecting an additional 177 variants (**Table S4**).

### Fine mapping analysis identifies three potential causal variants located in the *GATAD2A*, *SUGP1* and *MAU2* genes

We performed a fine mapping analysis incorporating all genome-wide significant SNPs to identify potential causal variants within each locus. This analysis revealed three candidate variants: rs57009615 (*GATAD2A* - intron 11), rs2240117 (*SUGP1 -* intron 3), and rs2285628 (*MAU2* - 3’ UTR). Each variant demonstrated strong evidence of a causal relationship with hepatic fat, as indicated by a PIP = 1.000 (**Table S5**). All three SNPs exhibited CADD scores suggesting a low likelihood of being deleterious; specifically, the scores were 6.705 for rs57009615, 0.284 for rs2240117, and 5.211 for rs2285628. In contrast, the RegulomeDB scores—indicative of the potential regulatory nature of these variants—varied significantly: 73.5% for rs57009615, 18.4% for rs2240117, and 60.9% for rs2285628. The *GATAD2A* variant was classified with a high rank of 2b (supporting data includes transcription factor binding along with any motif presence, footprint evidence, and chromatin accessibility peaks), whereas the *SUGP1* and *MAU2* variants had low ranks of 7 and 4 respectively. Overall, only rs57009615 demonstrated a high probability of being a relevant regulatory variant.

### MAGMA analysis identified *RFXANK,* a gene not detected in the GWAS

We performed MAGMA analyses, utilizing summary statistics from both the CC-GWAS and QT-GWAS. The CC-MAGMA analysis identified eight genes (total genes tested = 18,042; adj-p < 2.77E-06), all of which harbored variants with significant evidence of association in the CC-GWAS (**Fig S4A**, **Table S6A**). In the QT-MAGMA analysis, 19 genes were identified (total genes tested = 18,042; adj-p < 2.77E-06). Of these, only one gene, *RFXANK* (regulatory factor X associated ankyrin containing protein), represented a novel locus not previously detected in the QT-GWAS dataset (**Fig S4B** and **Table S6B**).

MAGMA gene set enrichment analysis did not reveal any significant GO functional classes (**Fig S5** and **Table S7**). The top functional classes for the CC-GWAS were “Srp dependent cotranslational protein targeting to membrane” (p = 7.9E-06, adj-p = 0.083) and “ATP binding cassette ABC transporter complex” (p = 9.3E-06, adj-p = 0.097). The top functional classes for the QT-GWAS were “Intraciliary transport involved in cilium assembly” (p = 8.9E-05; adj-p = 0.926) and “Very low-density lipoprotein particle assembly” (p = 1.1E-04; adj-p = 1.000).

### TWAS and fine mapping identify *SAMM50* as causal gene for hepatic steatosis

The TWAS analysis, utilizing liver data models from GTEx v8 to investigate the relationship between genetic variants and gene expression levels, did not reveal any new associations beyond those identified in the GWAS or MAGMA analyses. In the CC-TWAS, we identified one gene, *MBOAT7*, that remained significant after multiple testing correction (Z = −5.025; adj-p = 1.9E-03). This gene also passed joint and conditional testing, suggesting that its signal is independent of nearby SNPs (**Fig 4A** and **Table S8A**). While significant genetic variants in *MBOAT7* were not identified in the CC-GWAS, they were detected in the QT-GWAS and further confirmed in the QT-MAGMA analysis.

**Fig. 4.**
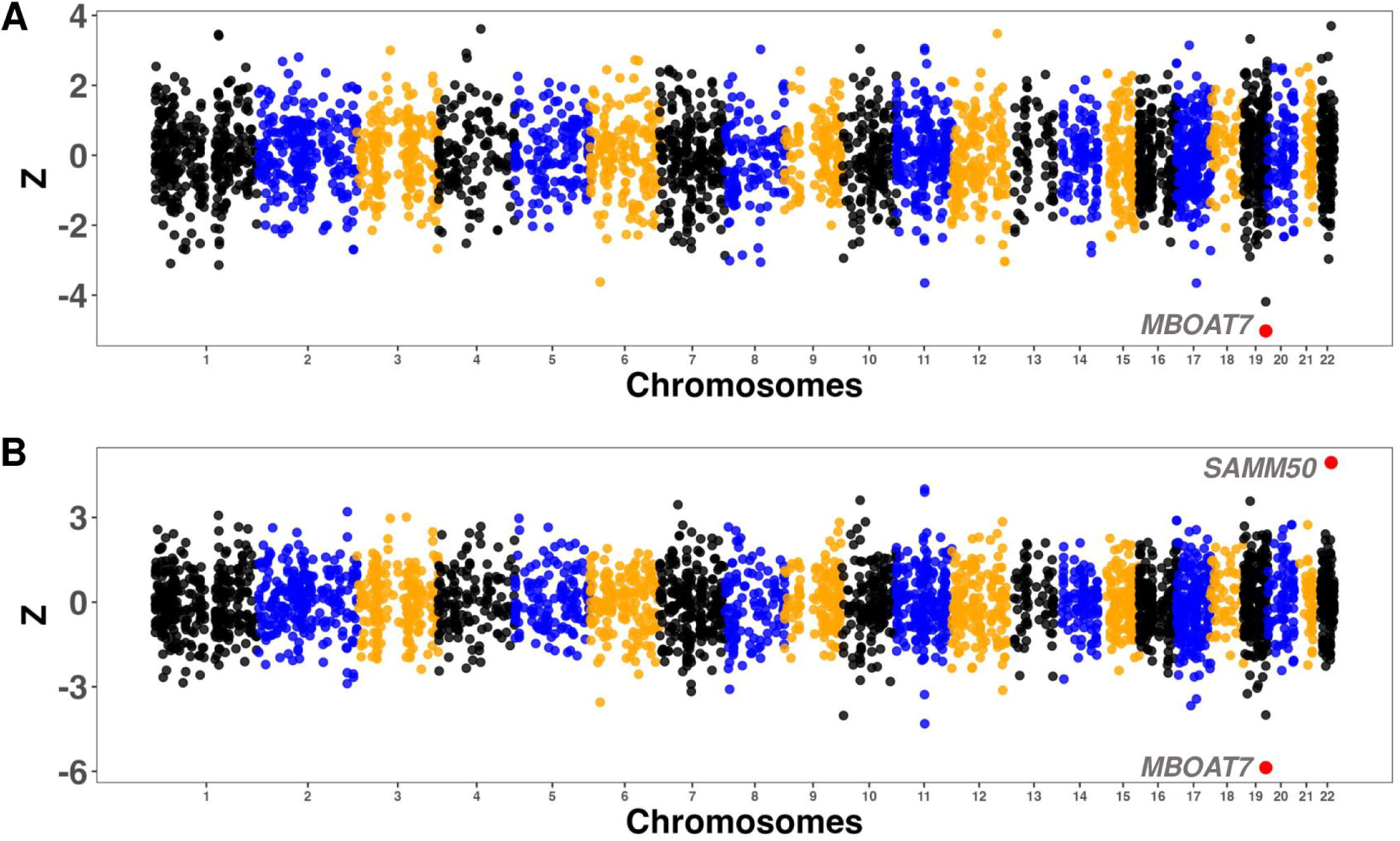
Miami plot illustrating the results of TWAS for the (A) CC-GWAS and (B) QT-GWAS. Genes exhibiting significant TWAS associations with liver fat, after applying Bonferroni correction accounting for the number of genes tested (n = 3,726; p < 1.34E-05) and passing the joint/conditional test are, are represented with red dots. Fine mapping analysis indicates strong evidence supporting *SAMM50* as a potential causal gene.

In the QT-TWAS, two genes, *MBOAT7* and *SAMM50,* emerged as significant, both passing the joint and conditional testing criteria (**Fig 4B** and **Table S8B**). Variants in the *SAMM50* gene were detected in the QT-GWAS (**Table S3**) and the QT-MAGMA analysis further substantiated the gene’s significance (**Table S6B**). TWAS fine mapping indicated a potential causal association with *SAMM50* (PIP = 1.000). In contrast, the evidence for *MBOAT7* was less definitive, with PIPs = 0.735 and 0.395 in the CC and QT analysis, respectively (**Table S8**).

We further conducted GSEA using the TWAS-Z as the effect size. Neither the CC nor QT analysis identified significantly enriched GO functional classes. However, the top processes overall included protein phosphatase binding (underexpressed), organellar large ribosomal subunit, and response to temperature stimulus (overexpressed) (**Table S9**).

### Comparison with previous GWAS in “lean NAFLD” identifies new variants

Two GWAS have previously investigated MASLD specifically in normal-weight individuals ^23,24^. One study, conducted in a Japanese population, did not achieve genome-wide significance for any SNPs after adjusting for sex and principal components ^23^. In contrast, the other study identified genome-wide significant signals at many loci ^24^. Our study overlapped with these findings, identifying 137 shared genetic variants from both the CC- and QT-GWAS (**Table S10** and **Table S11**). However, our CC-GWAS revealed 104 genetic variants not detected in the prior GWAS (**Table S12**), while the QT-GWAS identified 281 unique variants (**Table S13**). Some of these variants were found in *ARMC6* (armadillo repeat containing 6), *CILP2* (cartilage intermediate layer protein 2), *MAU2*, *MBOAT7*, *NR2C2AP* (nuclear receptor 2C2 associated protein), *PARVB* (parvin beta), *SLC25A42* (solute carrier family 25 member 42), *SUGP2*, *TM6SF2*, and *TMC4* (transmembrane channel like 4).

## Conclusions

Our study identified significant genetic associations on chromosomes 19 and 22 linked to hepatic steatosis in individuals with normal BMI. These findings challenge the prevailing view that genetic risk factors for MASLD primarily manifest in individuals with excess weight. The identification of variants in well-established genes such as *PNPLA3*, *SAMM50*, and *TM6SF2* emphasizes the role of genetic predisposition in the development of MASLD in normal weight individuals. These associations were robust across both CC-GWAS and QT-GWAS designs, further strengthening the relevance of these loci. Additionally, GWAS fine mapping identified potential causal variants in *GATAD2A*, *SUGP1* and *MAU2,* while TWAS fine mapping implicated *SAMM50* as a driver of steatosis in normal weight individuals, possibly through changes in gene expression. Our findings provide new insights into genetic determinants of steatotic liver in individuals who do not fit the typical clinical MASLD profile.

While interest in the genetic susceptibility to hepatic steatosis in nonobese individuals is not a new line of investigation, most previous studies have focused primarily on candidate genes, often yielding associations with modest effect size and limited genome-wide significance (^26^ and references therein). To date, only two studies have conducted genome-wide association investigations in normal weight populations. One of these studies, conducted in a Japanese population, did not find any variants reaching genome-wide significance ^23^. The sample size of 275 individuals with MASLD and 1,411 non-MASLD controls, may have had limited the power to detect genetic association. In contrast, the other study, which also utilized the UKB resource, identified genome-wide significant associations, including some that overlapped with the results reported here ^24^. When comparing our results with those the previous study ^24^, we observed association with variants in 11 genes, including *PNPLA3*, *SAMM50*, and *SUGP1*. However, we also identified new associations with genetic variants in *ARMC6*, *CILP2*, *MAU2*, *MBOAT7*, *NR2C2AP*, *PARVB*, *SLC25A42*, *SUGP2*, *TM6SF2*, and *TMC4* that were absent in the previous study. These novel associations could reflect differences in the specific populations studied, methodologies used, or the nature of hepatic steatosis in non-obese individuals. Surprisingly, we did not replicate the signal in the *HFE* gene, which had been the primary finding from the prior work. The lead variant in *HFE* identified in that study (rs1800562) was detected with p = 0.052 and p = 0.035 in our QT and CC-GWAS, respectively. The discrepancy in findings might be partially attributable to different sample selection criteria, as the previous GWAS utilized 12,804 controls, compared to the 10,103 in the current study. Unlike the prior work, we also assessed liver fat content as a quantitative trait, which strengthened the evidence of association, and performed several post-GWAS analyses that resulted in detection of potentially causal variants.

The *PNPLA3* SNP rs738409, the second strongest signal in our study, has been extensively linked to hepatic steatosis, steatohepatitis, fibrosis, cirrhosis, and hepatocellular carcinoma across diverse populations ^27–29^, particularly in individuals with obesity ^30–34^. The variant was also identified in the GWAS by Sun et al. ^24^ (p = 8.6E-13), suggesting a broader role in hepatic fat accumulation beyond obesity-related steatotic liver disease and independent of adiposity. The rs738409 variant is non-synonymous, leading to an isoleucine – to – methionine substitution at position 148 (I148M), which impairs triglyceride mobilization and promotes hepatic fat accumulation ^35^. The *TM6SF2* variant rs58542926, another well-documented variant associated with liver fat accumulation, similarly exhibits a role in hepatic steatosis in the absence of increased body mass.

*SAMM50* (Sorting and Assembly Machinery Component 50 Homolog) encodes a mitochondrial protein involved in maintaining mitochondrial morphology and promoting mitophagy, both of which are essential for mitigating the effects of oxidative stress in liver cells ^36,37^. The identification of *SAMM50* as a significant gene through our GWAS and TWAS analyses, along with its designation as a primary driver based on fine mapping, emphasizes the critical role of mitochondrial dysfunction and oxidative stress in the pathogenesis of liver fat accumulation in individuals with normal weight. Our findings are concordant with those of Li et al ^38^, who reported increased MASLD susceptibility in Chinese individuals with overweight (BMI > 23 kg/m^2^) carrying risk genotypes at rs738491 and rs2073082. Higher hepatic *SAMM50* transcript levels were observed in MASLD patients and lipid-loaded Hep3B cells ^38^. Interestingly, in vitro experiments demonstrated downregulation of *SAMM50* by rs738491 and rs2073082 variants, which resulted in impaired fatty acid oxidation and subsequent accumulation of lipid ^38^. Conversely, *SAMM50* overexpression was found to enhance fatty acid oxidation and reduce lipid buildup, suggesting that its deficiency directly contributes to the accumulation of lipids by limiting fatty acid breakdown. Our findings extend the association of *SAMM50* variants to normal-weight individuals, indicating that the role of SAMM50 in liver fat deposition is not solely dependent on obesity, and provides further evidence supporting a causal role for *SAMM50* in hepatic steatosis.

GWAS fine mapping revealed the presence of potential causal variants in *SUGP1*, *GATAD2A*, and *MAU2;* all these genes also emerged in the prior analysis ^24^. *SUGP1* (SURP And G-Patch Domain Containing 1) contributes to the regulation of cholesterol metabolism ^39^. An extended haplotype including rs10401969, significant in our CC-GWAS and QT-GWAS analyses, as well as the prior GWAS ^24^, has been linked with coronary artery disease, plasma low density lipoprotein cholesterol levels, and diverse energy metabolism phenotypes (^39^ and references therein). In addition, we previously observed a nominal association between rs10401969 and hepatic fat in individuals belonging to the severely obese BMI category (BMI > 40 kg/m^2^) (p = 3.1E−07) ^40^. While the functional consequences of rs10401969, an intronic variant, are unknown, some evidence suggests that it may impact alternative splicing ^39^ of the *SUGP1* transcript.

*GATAD2A* (GATA Zinc Finger Domain Containing 2A) encodes a protein that enables zinc ion binding. Low serum zinc levels have been observed in NAFLD ^41^. Zinc, whose homeostasis is primarily regulated in the liver, is associated with hepatic steatosis through various mechanisms, including antioxidant defense, insulin resistance, inflammation, and fibrogenesis ^42–44^. While zinc supplementation may reduce fibrosis levels, it had little effect on IHTG in mouse models of MASH ^45^. The potentially causal variant identified in our analysis, rs57009615, while having an intronic location, is predicted by RegulomeDB analysis to have regulatory and functional effects. Further studies may be warranted to explore the effect of this variant on *GATAD2A* RNA processing and expression.

*MAU2* (MAU2 Sister Chromatid Cohesion Factor), the remaining gene identified in our fine-mapping analysis, encodes a protein involved in sister chromatid cohesion during cell division. Currently, there is limited mechanistic evidence linking MAU2 to hepatic steatosis or metabolic dysfunction.

Our study has some limitations. First, while we included a substantial number of participants, the relatively small number of cases (n=815) compared to controls may affect the power to detect genetic associations, particularly those with a smaller effect size. However, our sample size is comparable to that of Sun et al ^24^, which identified significant genetic association in four loci, and an order of magnitude larger than the GWAS of Yoshida et al ^23^. Second, our cohort consists of individuals of European ancestry, which may limit the generalizability of our findings to other ethnic groups. Although many of the variants we detected have been observed in various populations, MASLD prevalence and genetic architecture vary across populations, necessitating validation in more diverse cohorts. Despite controlling for population stratification, some residual stratification may still affect our results.

Third, while our study identified significant associations and detected potential candidates for validation, the causal mechanisms behind these associations remain unclear, requiring functional validation. Prior work by Li et al ^38^ suggests that *SAMM50* variants reduce its expression, leading to impaired fatty acid oxidation and lipid accumulation, which may be relevant to our findings. Finally, MRI-PDFF was performed at a single time point, limiting our ability to capture the dynamic nature of liver fat accumulation or distinguish between different causes of hepatic steatosis. This could introduce heterogeneity and obscure associations specific to individuals with normal weight. Because this was a genetic analysis, the contribution of environmental or lifestyle influences such as diet and physical activity remained unconsidered, which could introduce residual confounding.

In conclusion, our study represents a significant step forward in understanding the genetic basis of hepatic steatosis in normal-weight individuals. The identification of novel genetic determinants, as well as the confirmation of previously detected associations, provides new insights into the pathophysiology of MASLD in this understudied population. As research into the genetic drivers of MASLD continues to evolve, our findings underscore the importance of considering genetic predisposition in the context of normal BMI, which may have distinct mechanisms and therapeutic needs compared to the broader MASLD population.

## Supporting information

Detailed methods

all Supplemental tables

## Data Availability

All data are available upon reasonable request to the authors.

## Abbreviations

BMI: body mass index
CC: case-control
eQTL: expression quantitative trait locus
IHTG: intrahepatic triglyceride
GATAD2A: GATA Zinc Finger Domain Containing 2A
GWAS: genome-wide association study
MAGMA: multi-marker analysis of GenoMic annotation
MASH: metabolic dysfunction-associated steatohepatitis
MASLD: metabolic-associated steatotic liver disease
MAU2: MAU2 Sister Chromatid Cohesion Factor
MBOAT7: Membrane-bound O-acyltransferase 7
MRI-PDFF: magnetic resonance imaging-proton density fat fraction
PIP: posterior inclusion probability
PNPLA3: patatin-like phospholipase domain-containing protein 3
QT: quantitative trait
SAMM50: Sorting and Assembly Machinery Component 50 Homolog
SNP: single nucleotide polymorphism
SUGP1: SURP And G-Patch Domain Containing 1
TWAS: transcriptome-wide association study
UKB: United Kingdom Biobank

## Acknowledgements

We acknowledge Drs. Mamatha Blat and Sareh Keshavarzi in guiding the cohort setup. This research was conducted using the UK Biobank Resource (Project ID 43036) and supported by the NIDDK (R01DK127015).

## Author contributions

ISP: data collection, data analysis and interpretation, drafting of manuscript; JD: data collection, data analysis and interpretation, critical review of the manuscript; NJS: critical review of the manuscript; JKD: conceptualization of the work, data interpretation, drafting of the manuscript, project administration, funding acquisition; final approval of the version to be published.

