## Supplementary material for "Potential causal links between genetic variants in *SAMM50*, *SUGP1*, *MAU2*, and *GATAD2A* and liver fat in individuals with normal weight": Detailed methods

### Supplementary Methods

Study sample. Data used in the following analyses was derived from the UKB database, which contains information from approximately 500,000 participants aged between 40 and 69 years recruited from the United Kingdom between 2006 and 2010 ^25^. The UKB database includes comprehensive health data, results from physical examinations, and biological samples for genetic analysis.

Data preprocessing, filtering, and quality control. An overview of the analytical workflow and study design is shown in **Fig 1**. The UKB imputed genotyped data version 3, encompassing 487,411 individuals and more than 96 million variants based on the GRCh37 genome build, was utilized for this study. Preprocessing, quality control, and genetic association analysis were performed using PLINK 2.0. Initially, we converted UKB bgen format files to PLINK bed/bim/fam format. During this conversion, we filtered the dataset to include only variants with high imputation quality, specifically utilizing the criterion of an ‘imputed information score (defined by UKB)’ > 0.8. Variants were restricted to those found on autosomal chromosomes. Consequently, this preprocessing step resulted in a dataset of 28,733,793 variants and 487,409 samples. Next, we applied quality control filters. Samples with heterozygosity values that were outside of three standard deviations were excluded, resulting in a sample size of 474,828 individuals. We also removed samples with a kinship value ≥ 0.125 based on UKB genetic relatedness data. Individuals who had withdrawn their consent up until the last update (April 25, 2023) prior to the analysis were excluded. To ensure consistency, we cross-referenced the UKB genetic sex (UKB data field 22001) and self-reported sex (UKB data field 31) and retained samples with consistent sex information across both fields. To establish a homogenous ethnic group and minimize confounding effects stemming from ancestry and environmental factors, we selected individuals with a self-reported ancestral background of ‘White British’ (UKB data field 21000) and refined the selection by choosing ‘Caucasians’ from the genetic ethnic grouping (UKB data field 22006), which constitutes the largest single ethnic group comprising > 80% of the samples. This step also involved removing outliers based on genetic principal components. In addition to these filters, we restricted the sample to individuals with unambiguous values for the phenotype and covariates used in the subsequent association analysis. For the current study, we selected individuals from the UKB database with available MRI-PDFF values (recorded in UKB field 40061-2.0) up until the date of the analysis, when 40,532 participants from the entire UKB cohort had these measurements. From this group, only participants who met the quality control criteria described above were selected. To exclude additional potential confounding factors, we identified participants with conditions that might impact liver fat levels using ICD9 and ICD 10 codes within the UKB diagnostic and death records (**Table S1**) and removed them from the selected group. When we limited selection to participants classified within the normal BMI category, defined as BMI < 25 kg/m^2^, we achieved a sample size of 10,934 individuals.

For the association analyses described in the next section, we performed additional filtering using specific PLINK parameters. These included a minor allele frequency of 0.05, missingness per individual of 0.05, missingness per marker of 0.05, and Hardy-Weinberg equilibrium of 1E-6. The application of these filtering criteria resulted in a reduction of the sample size to 10,918 individuals and a total of 6,156,170 variants. The principal component analysis plot utilizing this final dataset is shown in **Fig S1**.

Genetic association analysis. We conducted two distinct GWAS. The first analysis utilized a case-control study design (CC), where MRI-PDFF values were used to define individuals with or without hepatic steatosis. The second analysis was a quantitative study (QT), directly using the range of MRI-PDFF values. Both studies employed the same filtering parameters, as described in the previous section, and utilized the same set of covariates, including birth year, sex, BMI, alcohol intake frequency (UKB field 1558), and the first 10 genetic principal components (UKB field 22009). To ensure consistency and comparability, we standardized each covariate using the 'covar-variance-standardize' function in PLINK ^26^. For the association analyses, we employed the 'glm' function in PLINK, allowing us to assess the genetic associations while accounting for the specified covariates.

For the CC study (CC-GWAS), we defined hepatic steatosis as MRI-PDFF ≥5% (cases) and normal liver as MRI-PDFF <5% (controls). Using PLINK, we calculated odds ratios (OR) and P-values for each variant, adjusting for the specified covariates. To account for multiple testing, we applied the established genome-wide cutoff of p < 5.0E-08. To analyze MRI-PDFF in the QT analysis (QT-GWAS), we used the same ‘glm’ function in PLINK, but with continuous values of MRI-PDFF in the phenotype field. In this case, the linear model provided effect values (beta) and P-values for each variant. We used the same parameters and covariates as in the CC-GWAS analysis and accounted for multiple testing using the same cutoff. Following both analyses, we employed the ‘clump’ function in PLINK 1.9 to extract variants with a P-value < 5.0E-08 and removed variants that were in linkage disequilibrium (LD) with the most significant variants. The clumping parameters used were clump-p1 = 5.0E-08, clump-r2 = 0.1, clump-kb = 250. SNPs were annotated with their corresponding genes utilizing the ANNOVAR software tool, referencing the human genome assembly version hg19.

GWAS fine-mapping. We conducted a fine-mapping analysis with the goal of identifying the causal variants. The analysis was performed using the *FINEMAP v.1.4* software, which employs an algorithm that explores a set of the most probable causal configurations of the region. For both the CC and QT analyses, we selected all SNPs significant at the genome-wide level of p < 5.0E-08. The LD correlation matrix was estimated using *PLINK2* with the “--r square” function. Fine mapping was carried out using the *stochastic statistic search* method, considering the value of a posterior inclusion probability (PIP) > 80% as strong evidence of causality, a PIP ≥ 50% as moderate evidence of causality, and a PIP < 50% as weak evidence of causal association between the variant and the trait. We also utilized the Log_10_ Bayes Factor (LBF), which indicates the strength of association between a variant and the trait of interest without inferring causality. A LBF ≥ 2 implies strong evidence of association, while 1 < LBF ≤ 2 indicates moderate evidence for association, 0.5 < LBF ≤ 1 indicates suggestive evidence, and LBF ≤ 0.5 indicates weak evidence of association.

MAGMA. GWAS summary statistics were analyzed with the Multi-marker Analysis of GenoMic Annotation (MAGMA) method, which provides gene-level statistics using a multiple regression approach to incorporate LD information between markers and detect multi-marker effects. The statistic was generated considering 10 kb regions surrounding the gene boundaries (parameter *--annotate window=10,10*). P-values were adjusted for multiple testing using the Bonferroni method, accounting for the number of genes tested. Results were investigated for Gene Ontology enrichment through the MAGMA gene-set analysis.

TWAS. We utilized liver expression data models from the Genotype-Tissue Expression Project version 8 (GTEx v8) –European reference data – to impute gene expression values. These values quantify the relationship between individual genotype and corresponding gene expression levels, capturing *cis*-acting genetic effects. We matched genetic variants using “rsID” rather than genomic position to avoid issues arising from differences in genomic coordinates between GTEx v8 (GrCh38) and the GWAS dataset. Using GTEx v8 data, we computed gene weights to estimate the effects of individual SNPs on gene expression, which were then used to evaluate whether the predicted levels were associated with the phenotype. TWAS p-values were adjusted using the Bonferroni method based on the total number of genes included in the GTEx v8 liver reference (n = 3,726; p < 1.34e-05; alpha = 0.05). Joint and conditional tests were conducted for all genes with suggestive adjusted p-value (p < 2.68e-05; alpha = 0.10), to assess whether the signal in genome-wide significant genes was independent of variants located in nearby loci. Genes that exhibit joint significance are independent of variants located in adjacent loci, while genes classified as conditionally non-significant lose significance upon adjustment for the contributions of nearby genes. To identify causal genes associated with the trait, we performed TWAS using the FOCUS method, which allows derivation of the covariance structure among TWAS statistics by integrating the LD and eQTL weights utilized in the prediction. The TWAS association statistics for all genes within a risk region were modeled using a multivariate Gaussian distribution, which is parameterized by effect sizes at causal genes, residual SNP effects, and the correlation structure induced by inferred expression weights in conjunction with LD. To mitigate potential bias arising from pleiotropic effects of SNPs, we incorporated an intercept term that quantifies the average SNP effect sizes associated with predicted expression.
